## Supplementary material for "Investigation and public health response to a COVID-19 outbreak in a rural resort community — Blaine County, Idaho, 2020": S3 Figure

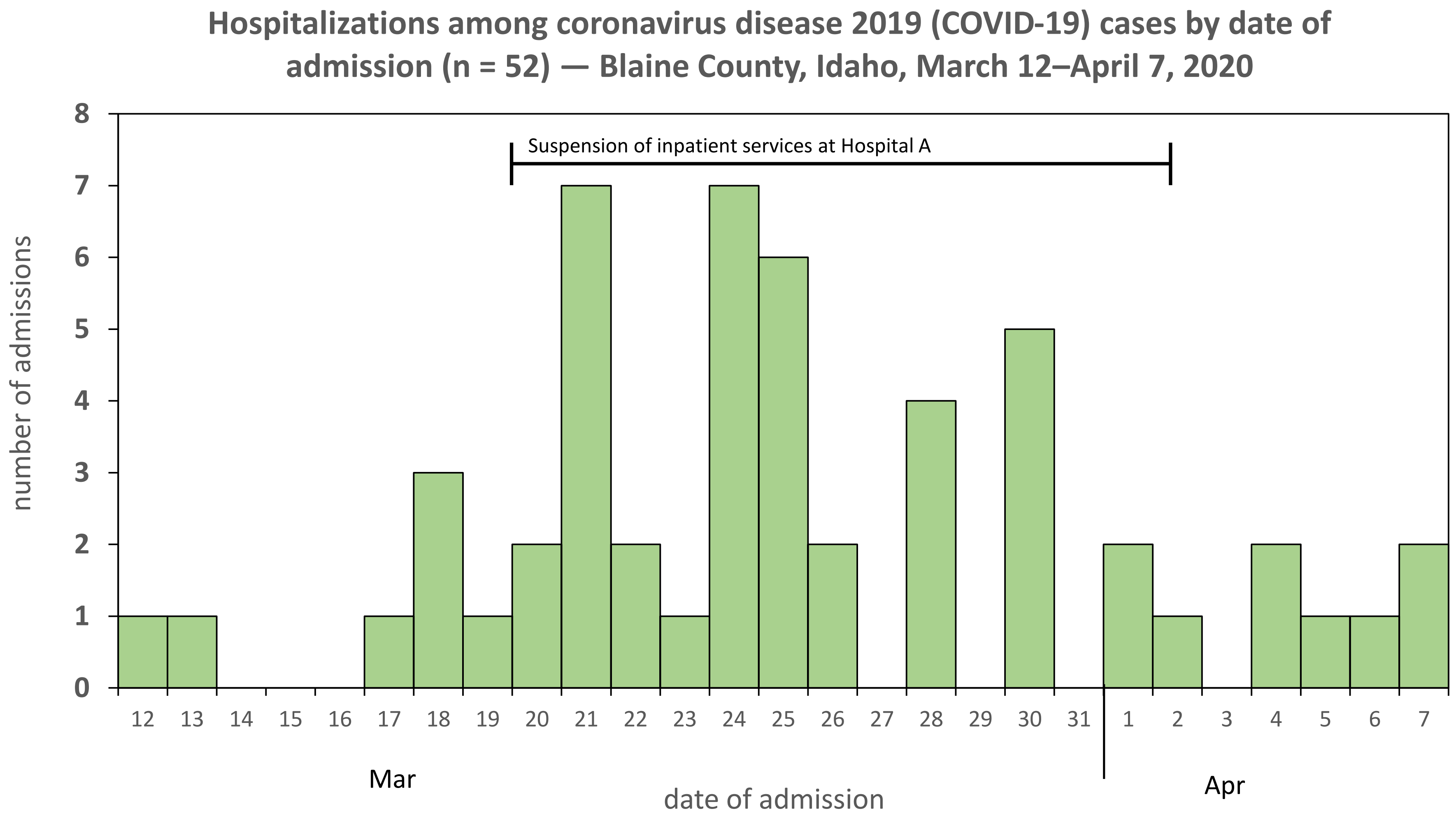


**S3 Figure.** Hospitalization of Blaine County residents for COVID-19 by date of admission. Admission dates from cases reported from March 13–April 10 are included on the graph.
