## Supplementary material for "Investigation and public health response to a COVID-19 outbreak in a rural resort community — Blaine County, Idaho, 2020": S4 Figure

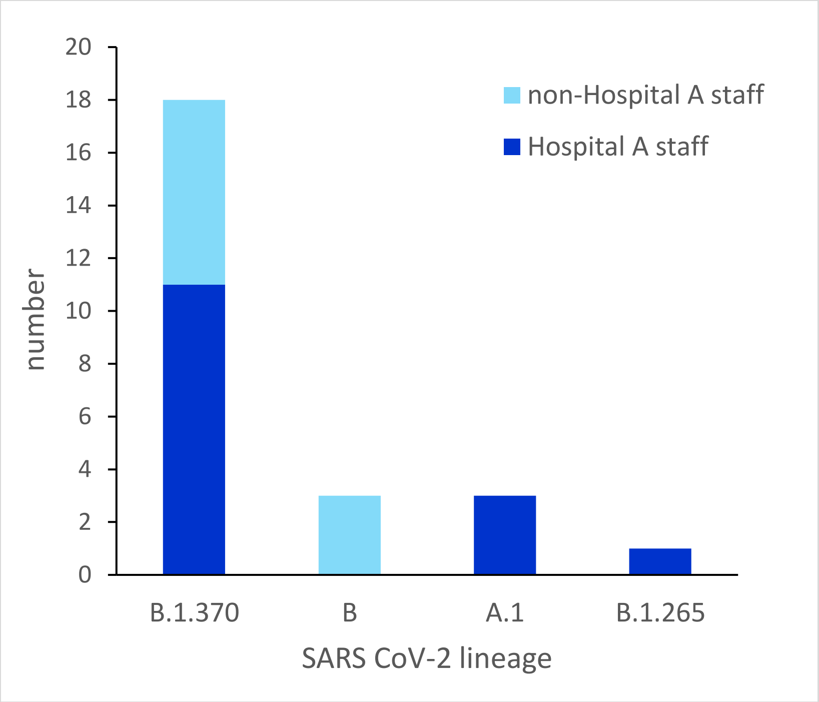


**S4 Figure.** SARS-CoV-2 lineages identified from Blaine County residents who work at Hospital A (Hospital A staff, dark blue) and Blaine County residents who do not work at Hospital A (non-Hospital A staff, light blue).
