## Supplementary material for "Investigation and public health response to a COVID-19 outbreak in a rural resort community — Blaine County, Idaho, 2020": S5 Table

**S5 Table. Distribution of confirmed COVID-19 cases (n = 402) and census block group characteristics for Blaine County, Idaho.** Census block group data are 2014-2018 American Community Survey 5-year estimates.

| | | Census Block Group Characteristics | | | | | | |
|---|---|---|---|---|---|---|---|---|
| | | | | | *Median household income (USD)* | | | |
| **Census block group** | **COVID-19 cases (%)** | **Population** | **Hispanic or Latino origin** | **Persons in renter-occupied housing** | **under $25K** | **$25K to $75K** | **$75K to $150K** | **over $150K** |
| Block Group 1, Census Tract 9601 | 3 (0.7) | 1304 | 12.9% | 40.8% | 25.5% | 39.3% | 25.2% | 10.0% |
| Block Group 2, Census Tract 9601 | 11 (2.7) | 1421 | 15.6% | 37.2% | 22.3% | 52.7% | 18.3% | 6.8% |
| Block Group 3, Census Tract 9601 | 23 (5.7) | 1478 | 4.1% | 13.7% | 18.0% | 42.8% | 31.5% | 7.6% |
| Block Group 4, Census Tract 9601 | 14 (3.5) | 571 | 29.8% | 37.2% | 29.5% | 41.5% | 29.0% | 0.0% |
| Block Group 1, Census Tract 9602 | 37 (9.2) | 1313 | 20.0% | 54.1% | 46.4% | 39.0% | 8.3% | 6.4% |
| Block Group 2, Census Tract 9602 | 44 (10.9) | 2667 | 6.5% | 24.3% | 15.3% | 28.8% | 32.8% | 23.1% |
| Block Group 3, Census Tract 9602 | 70 (17.4) | 5052 | 47.1% | 49.2% | 19.3% | 49.5% | 25.9% | 5.4% |
| Block Group 4, Census Tract 9602 | 15 (3.7) | 1346 | 41.8% | 13.5% | 6.5% | 56.4% | 32.4% | 4.7% |
| Block Group 1, Census Tract 9603 | 48 (11.9) | 1043 | 4.6% | 11.1% | 15.4% | 37.6% | 33.4% | 13.6% |
| Block Group 2, Census Tract 9603 | 19 (4.7) | 878 | 14.4% | 33.0% | 27.9% | 35.8% | 29.3% | 7.0% |
| Block Group 3, Census Tract 9603 | 54 (13.4) | 1800 | 10.9% | 49.0% | 13.0% | 50.2% | 30.8% | 6.0% |
| Block Group 1, Census Tract 9605 | 32 (8.0) | 1835 | 21.2% | 25.8% | 27.4% | 28.8% | 32.7% | 11.1% |
| Block Group 2, Census Tract 9605 | 32 (8.0) | 1286 | 3.3% | 21.7% | 20.1% | 45.2% | 23.5% | 11.2% |
