## Supplementary material for "Investigation and public health response to a COVID-19 outbreak in a rural resort community — Blaine County, Idaho, 2020": S1 Table

**GISAID Idaho Sequence Key**

hCoV-19/USA/ID-UNM-IBL021/2020
hCoV-19/USA/ID-UNM-IBL022/2020
hCoV-19/USA/ID-UNM-IBL023/2020
hCoV-19/USA/ID-UNM-IBL025/2020
hCoV-19/USA/ID-UNM-IBL026/2020
hCoV-19/USA/ID-UNM-IBL028/2020
hCoV-19/USA/ID-UNM-IBL030/2020
hCoV-19/USA/ID-UNM-IBL031/2020
hCoV-19/USA/ID-UNM-IBL032/2020
hCoV-19/USA/ID-UNM-IBL033/2020
hCoV-19/USA/ID-UNM-IBL037/2020
hCoV-19/USA/ID-UNM-IBL038/2020
hCoV-19/USA/ID-UNM-IBL039/2020
hCoV-19/USA/ID-UNM-IBL040/2020
hCoV-19/USA/ID-UNM-IBL041/2020
hCoV-19/USA/ID-UNM-IBL052/2020
hCoV-19/USA/ID-UNM-IBL053/2020
hCoV-19/USA/ID-UNM-IBL055/2020
hCoV-19/USA/ID-UNM-IBL056/2020
hCoV-19/USA/ID-UNM-IBL058/2020
hCoV-19/USA/ID-UNM-IBL061/2020
hCoV-19/USA/ID-UNM-IBL062/2020
hCoV-19/USA/ID-UNM-IBL063/2020
hCoV-19/USA/ID-UNM-IBL067/2020
hCoV-19/USA/ID-UNM-IBL068/2020
hCoV-19/USA/ID-UNM-IBL150/2020
hCoV-19/USA/ID-UNM-IBL154/2020
hCoV-19/USA/ID-UNM-IBL208/2020
hCoV-19/USA/ID-UNM-IBL209/2020
hCoV-19/USA/ID-UNM-IBL217/2020
hCoV-19/USA/ID-UNM-IBL273/2020
hCoV-19/USA/ID-UNM-IBL311/2020
hCoV-19/USA/ID-UNM-IBL312/2020
hCoV-19/USA/ID-UNM-IBL359/2020

**GISAID US and Type Sequence Key**

NC_045512.2 SARS-CoV-2 isolate Wuhan-Hu-1
hCoV-19/USA/AZ-TG266871/2020
hCoV-19/USA/CA-CZB-9705/2020
hCoV-19/USA/CA-SCCPHD-UC122/2020
hCoV-19/USA/CA-SCCPHD-UC130/2020
hCoV-19/USA/LA-CDC-0499/2020
hCoV-19/USA/MS-CDC-6567/2020
hCoV-19/USA/NY-NYCPHL-000054/2020
hCoV-19/USA/NY-NYCPHL-000356/2020
hCoV-19/USA/NY-NYCPHL-000591/2020
hCoV-19/USA/NY-NYCPHL-000607/2020
hCoV-19/USA/NY-Wadsworth-11017-01/2020
hCoV-19/USA/NY-Wadsworth-11249-01/2020
hCoV-19/USA/NY-Wadsworth-12437-01/2020
hCoV-19/USA/RI_0882/2020
hCoV-19/USA/VA-DCLS-0018/2020
hCoV-19/USA/WA-UW119/2020
hCoV-19/USA/WA-UW-192/2020
hCoV-19/USA/WA-UW-195/2020
hCoV-19/USA/WA-UW71/2020
